# Adjusted Dynamics of COVID-19 Pandemic due to Herd Immunity in Bangladesh

**DOI:** 10.1101/2020.09.03.20186957

**Authors:** Md. Enamul Hoque, Md. Shariful Islam, Mohammad Ruhul Amin, Susanta Kumar Das, Dipak Kumar Mitra

## Abstract

Amid growing debate between scientists and policymakers on the trade-off between public safety and reviving economy during the COVID-19 pandemic, the government of Bangladesh decided to relax the countrywide lockdown restrictions from the beginning of June 2020. Instead, the Ministry of Public Affairs officials have declared some parts of the capital city and a few other districts as red zones or high-risk areas based on the number of people infected in the late June 2020. Nonetheless, the COVID-19 infection rate had been increasing in almost every other part of the country. Ironically, rather than ensuring rapid tests and isolation of COVID-19 patients, from the beginning of July 2020, the Directorate General of Health Services restrained the maximum number of tests per laboratory. Thus, the health experts have raised the question of whether the government is heading toward achieving herd immunity instead of containing the COVID-19 pandemic. In this article, the dynamics of the pandemic due to SARS-CoV-2 in Bangladesh are analyzed with the SIRD model. We demonstrate that the herd immunity threshold can be reduced to 31% than that of 60% by considering age group cluster analysis resulting in a total of 53.0 million susceptible populations. With the data of Covid-19 cases till *July* 22, 2020, the time-varying reproduction numbers are used to explain the nature of the pandemic. Based on the estimations of active, severe, and critical cases, we discuss a set of policy recommendations to improve the current pandemic control methods in Bangladesh.

## 1 Introduction

In late December 2019, a novel strain of Coronavirus (COVID-19) was first identified in Wuhan, a city in central China and thought to spread an outbreak of viral pneumonia [1, 2]. This novel Coronavirus is assumed to have a zoonotic origin, as all the very first pneumonia cases with unknown etiologies were linked with Wuhan’s Huanan Seafood Wholesale market [3]. Since the novel pathogen rapidly transmitted across the globe, the World Health Organization (WHO) officially declared the outbreak of COVID-19 to be a public health emergency of international concern on January 30, 2020 [4]. It has been reported by WHO that this contagious disease control heavily relies on early detection, isolation of symptomatic cases, prompt treatment, and tracing the contacts history [5]. However, the lack of antiviral vaccines and the difficulties in the identification of the asymptomatic carriers made it very challenging to contain the pandemic [6].

To examine the course of the pandemic and to suggest preventive strategies, mathematical models can play a crucial role in describing transmission dynamics as well as for forecasting quantitative assessment. The classic mean-field Susceptible-Infected-Recovery-Death (SIRD) model by Kermack and McKendrick is one of the most commonly used models that illustrates the quantitative pictures of a pandemic through four mutually exclusive infection phases, namely susceptible, infected, recovered and death [7, 8, 9, 10, 11]. Although several other mathematical models, including discrete-time SIR model [12], SEIR (susceptible, exposed, infected, recovered) model [13], control-oriented SIR model [14] and SIDARTHE (susceptible, infected, diagnosed, ailing, recognized, threatened, healed, extinct) models have been utilized so far to portray the progressive spread of the COVID-19 pandemic [15], we implemented SIRD model to explore the temporal progression of COVID-19 transmission in Bangladesh.

On March 26, 2020, a nation-wide 10-day lockdown was imposed by the Bangladesh government to contain the outbreak of COVID-19 in the wake of four deaths and at least 39 Corona active cases [16]. While there are many pivotal lessons to be learned from the US, Italy, Spain, China and other countries concerning their COVID-19 responses, it has been an uphill battle to import all their policies, such as prolonged lockdown, massive Coronavirus testing, isolation of positive cases and access to proper healthcare in Bangladesh due to its socio-economic structure. Despite nationwide lockdown, Bangladesh had seen a rise in the COVID-19 positive cases in the following few months.

On May 31, 2020, Bangladesh had a total of 47,153 positive cases, with a total death toll of 650. Despite the rise in the transmission rate, the government of Bangladesh decided to relax the lockdown restrictions to restore the economy in the country. As a preparation to face the pandemic, the Directorate General of Health Services in Bangladesh (DGHS) had prepared a total of 7,034 hospital beds that were outnumbered by the current need of the densely populated country [17].

Since the hospitals are usually centralized in Dhaka, the capital of Bangladesh, the outcry for hospital beds for the COVID-19 patients could be visible via countless news stories.

The total COVID-19 positive patients in Bangladesh has been reported to be a total of 213,254, with a total case fatality of 2,751 persons throughout the country on *July* 22, 2020. Regardless of lifting the lockdown, the DGHS restricted the number of lab tests which is quite contrary to the test and isolation policy. The result is clearly visible in Figure 1. As the number of lab tests has been decreasing, the number of daily positive cases started dropping as well. On the other hand, we observe that the test positivity rate is on the rise. In addition to this, there has been news in the daily newspapers that people are waiting for COVID-19 tests in a long line and many of them are returning without the tests due to the limited number of test kits to handle a large number patients [19]. Moreover, having a limited number of hospital beds together with unavailability of Oxygen supply, many patients stopped seeking medical help. As a result, while in the middle of June, national dailies reported that patients are dying without any medical help on the road while moving from one hospital to another for an empty bed [20], recently a lot of COVID-19 patients stopped going to the hospital to seek medical help resulting in empty beds [17].

**Fig. 1.**
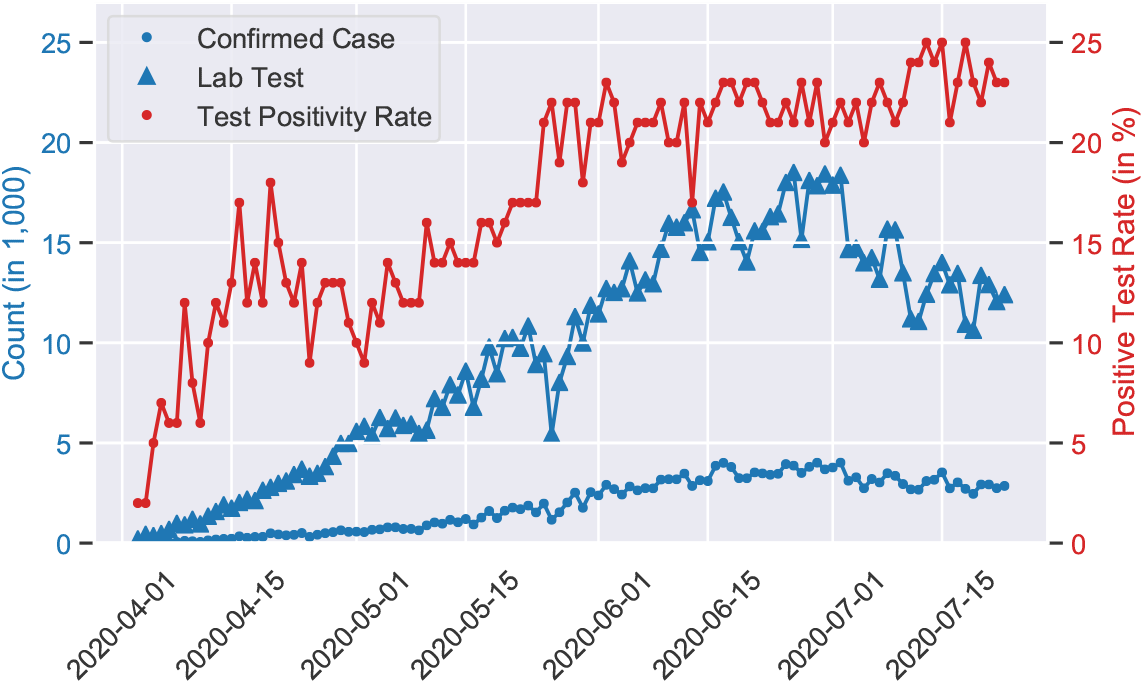
Bangladesh has the lowest number of tests per million people in the world. Still, a significant reduction in the RT-PCR based lab tests has been observed from July 1, 2020, in Bangladesh (blue triangle). This results in an increase in the test positivity rate approaching to 25% (red dot) within the following two weeks. WHO recommends that the test positivity rate should be less than 5% [18].

Therefore, based on the current situation, one can ask if Bangladesh is moving toward herd immunity and if so then what could be its consequence on the public health. It is indispensable to have a careful balance between various key epidemiological factors in order to estimate the spread of COVID-19 in Bangladesh. This is the reason why mathematical estimation can play a very important role to understand the future we may expect based on the latest policies implemented in the country. In the following sections of this article, we present the dynamics of the COVID-19 pandemic in Bangladesh with the SIRD model and estimate the number of susceptible populations as well as case fatalities rate to achieve the herd immunity in Bangladesh.

## 2 The Study of COVID-19 Pandemic in Bangladesh

Bangladesh has implemented various types of control measures to contain the pandemic, such as total/partial lockdown, declaring an area as a quarantined/red zone, suspending the long and short distance transportation in the different parts of the country, including wearing masks and maintaining social distance in the public transportation. Each of these control measures were imposed on various dates and were lifted off for various reasons. To describe the effect of the implemented pandemic control methods in Bangladesh, the time-varying reproduction number, *R_t_* is used since it provides the real-time estimation of the pandemic dynamics. In the case of controlling the pandemic of a region, the *R_t_* provides valuable information about the dynamics and instantaneous effect of the control mechanism [26]. We analyze the dynamics of SARS-CoV-2 cases in Bangladesh using *R_t_* with respect to four of the major events, i.e., starting of lockdown, mass movement, ease of lockdown (limited shopping mall, public transportation etc.), and lifting of the lockdown for restarting local economy.

The pandemic control method and the changes in SARS-CoV-2 cases are shown in Figure 2. Bangladesh implemented very early lockdown on March 26, 2020 to control the SARS-CoV-2 pandemic [22], and that was lifted on May 30, 2020 with 44,608 confirmed cases and 610 deaths [25]. In this report, we choose 100 confirmed cases as the baseline of *R_t_* computation that had climbed to 4.1 and then fell below 1.5 with high confidence on April 24, 2020.

**Fig. 2.**
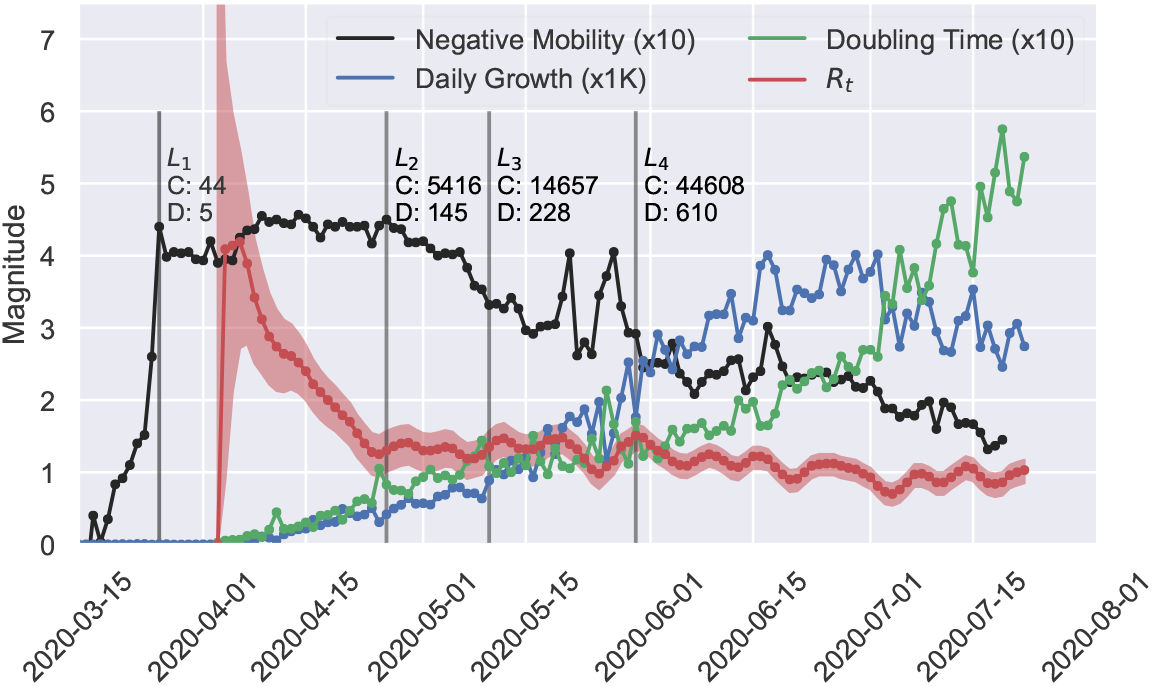
The time-varying reproduction number *R_t_*, has been compared with the average mobility [21], the growth of confirmed cases and the doubling time to analyze the effect of the implemented lockdown (*L*_1_ to *L*_4_) in Bangladesh. The lockdown situations are described as *L*_1_, March 26 when the lockdown is implemented for all over the country [22]; *L*_2_, April 26 when ready made garment factories reopened [23]; *L*_3_, May 10 when shopping malls were reopened before Eid-ul-Fitre [24]; and *L*_4_, May 30 when everything but educational institutes were reopened [25]. The confirmed cases and the number of deaths on those days are also shown (labeled as C and D).

Due to the garment factories reopening on April 26, 2020, a mass population moved to the capital of Bangladesh (*L*_2_) [23]. Despite of this movement, the value of *R_t_* remained around 1.25. During *L*_3_, all the shopping mall, business office, public transportation were reopened and surprisingly there was not any noticeable variation in *R_t_* [24]. We further observe that *R_t_* had dropped below 1.0 on May 25 and then it jumped to 1.5 on May 30 (*L*_4_), the day when Bangladesh reopened its economics by unlocking everything except the educational institutes [25].

Since the lifting of the lockdown (*L*_4_), we observe that public mobility is on the rise, represented as a drop in the negative Google mobility score in the Figure 2. That directly contradicts the drop in the value *R_t_*, drop in the daily growth rate and increase in the doubling time (Figure 2). In addition to these countering observations, in the Figure 1, we present that the test positivity rate is on the rise. Thus, it clearly present that the most successful intervention of test and isolation for controlling the COVID-19 pandemic has not been carefully maintained by the DGHS in Bangladesh. This is why, a discussion is surfacing among the public health professionals as though Bangladesh government is expecting to achieve herd immunity [27].

## 3 Adjustment of COVID-19 Dynamics in Herd Immunity

In this article, the dynamic behavior of the class of infected people is described using three different reproduction numbers, such as the basic (*R*_0_), effective (*R_e_*), and time-varying (*R_t_*) reproduction number. To estimate the number of people to be infected in case of the herd immunity, we consider the total number of current population to be 170.1 million by 2020 [28]. According to the SIR model, *R*_0_ is used to describe the transmissibility of a disease in a region if the population is randomly mixed. It will have only one value with a marginal error for an epidemic. But with the implementation of the various epidemic control methods in Bangladesh, the “randomly mixed” condition for virus transmission within the population has been violated. Currently, a few different *R*_0_ estimations have been reported ranging between 1.4 to 3.0 [29, 30, 31] for the country. But due to the intervention mechanism, the original method to compute susceptible population from the *R*_0_ cannot be followed. So we rather estimate the susceptible population for the herd immunity based on the current transimissiblity of COVID-19 within the individual age group.

**Table 1.** The age group distribution of the SARS-COV-2 cases in Bangladesh where cumulative confirmed and death cases were 74,865 and 1,012, respectively on *July* 22, 2020 [16, 28].

| Age Group (in years) | 1 - 20 | 21 - 40 | 41 - 60 | 60+ |
| --- | --- | --- | --- | --- |
| Population (%) | 43.5 | 32.6 | 16.5 | 7.5 |
| Confirmed Cases (%) | 10.2 | 54.6 | 28.4 | 6.7 |
| Deaths (%) | 2.86 | 11.57 | 48.49 | 37.08 |

Bangladesh has already implemented various types of control measures to contain the pandemic. Each of these control measures was imposed on various dates and was lifted off for various reasons. These random events made it very challenging to estimate the susceptible population for herd immunity in Bangladesh. Hence, we consider the initial susceptible population following the age group distribution of the population depending on the confirmed positive cases to make our final assessment for herd immunity. In the Table 1, we present the age distribution of the population in Bangladesh (collected from Socioeconomic Data and Application Center, or SEDAC) along with the confirmed positive cases and deaths for each of those age groups (collected from Institute of Epidemiology, Disease Control and Research, or IEDCR). In Bangladesh, the gender ratio for the confirmed cases has been reported as *Male*: *F emale* = 71: 29 by the IEDCR [16]. Using these distributions of population and the confirmed SARS-COV-2 cases, the susceptible population for herd immunity is estimated. As 54.6% of the total confirmed positive cases has been reported within the age group of *21–40*, we estimate the total susceptible population with respect to this age group.

### 3.1 Adjustment of Susceptible Population

We consider the population of Bangladesh is *P*, distributed in the age group as,

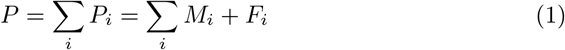

where *P_i_* is the population at *i*-th age group which is redistributed in male (*M*) and female (*F*).

Let us assume that *q*% of the male at age group *j* will be infected i.e., the susceptible population at *j, S_j_* = *M_j_ × q*%. In our case, we consider the age group *j* to be 21 *−* 40.

From the age distribution of the SARS-CoV-2, we can find the distributed number of all confirmed cases *C* as,

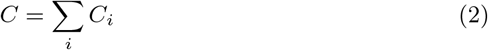

where *C_i_* is the confirmed cases at *i*-th age group, where the ratio of male (*CM*) and female (*CF*) is *m*: *f*. Thus,

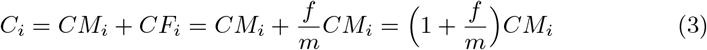

Since, the age group *j* (21–40 in Table 1) is the mostly affected group in Bangladesh due to their activities and movement for earning livelihood, we consider that this scenario of higher transmissibility of COVID-19 in the same age group will persist. We assume that rest of the population that will be affected in herd immunity can be estimated in proportion to the people affected in the age group *j*. So to find the susceptible population with respect to the age group *j*, we compute the proportion, *x* = *S_j_/CM_j_*. This leads us to approximate the number of susceptible population as,

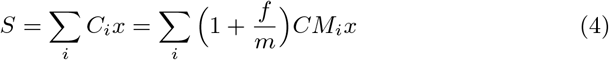

**Table 2.**
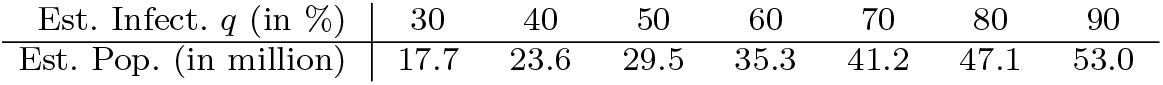
Estimation of the population to be infected by SARS-COV-2 in Bangladesh with respect to the baseline age group *21–40*.

The Table 2 shows the approximated susceptible population of Bangladesh for various rate of infection with respect to the age group 21–40. Therefore, if 30% of the total working population in Bangladesh get affected by COVID-19 to achieve herd immunity, we estimate that a total number of positive cases would be approximately 17.7 million. Similarly, if 90% of the population at the age group of *21–40* get affected, we would observe a total 53 million positive cases in Bangladesh. If we consider that 90% of the population at the age group of 21–40, only 31.1% of the population (53.0 of 170.1 million) of Bangladesh required to be infected by SARS-COV-2 to be in a state of herd immunity (Figure 3).

**Fig. 3.**
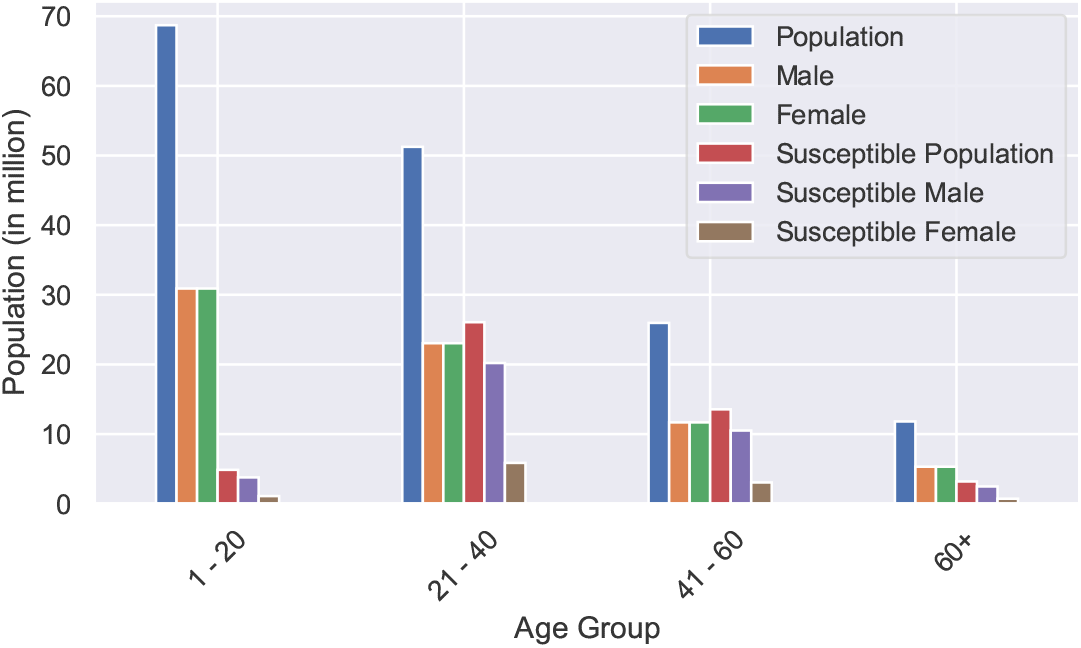
The age group based population distribution, along with gender, in Bangladesh has been used to estimate the initial susceptible population for SIRD model. To achieve the Herd Immunity of the people of Bangladesh, 53.0M of infected population has been considered at the age group of 21–40 years. This has been extrapolated in other age group with a distribution of confirmed case of Bangladesh [16]. The estimated distribution of initial susceptible population, along with gender, is also shown.

The above computation indicates that the herd immunity threshold can be reduced from 60% to 31% by categorizing the population into age groups. There is another major reason why we may observe such a low threshold in case of herd immunity. Due to the closure of all educational institutions from the very early stage of this pandemic in Bangladesh, the population in the age group 1–20 will not be the primary agent for spreading the virus. Around 43% of the total population belong to this age group, hence reducing the herd immunity threshold.

### 3.2 Dynamics of COVID-19 Cases In Herd Immunity

To estimate the dynamics of the COVID-19 cases, such as confirmed, recoverd, and death cases, in case of herd immunity, we have used the Unscented Kalman Filter (UKF). We are explaining the reason why UKF is very important to derive the dynamics. The differential equations in the SIRD model computes the values for an instantaneous event. Whereas, the COVID-19 cases that we observe each day is not instantaneous rather a discrete event on a wider time range. The events of a person actually being affected, detected, and reported of COVID-19 happen in different times, and thus loose the required instantaneous effect in the differential equation based SIRD model. Hence, we use the prediction based UKF model to derive the dynamics. The Jacobian matrix for designing the UKF algorithm is presented in the Supplementary Section (Equation 13). The uncertainty associated with the inference is estimated using the Merwe Sigma Points that deal with the hidden states of the considered system in an optimal and consistent way for a set of noisy and/or incomplete observation.

The dynamics of the estimated active cases for both the scenarios considering 53.0 million and 17.7 million initial susceptible population is shown in Figure 4. It had been reported that the country has the hospital facility of 7,034 SARS-COV-2 patients [17]. In a guideline, provided by the Health Ministry of Bangladesh [32], around 16% of the active cases may need hospitalization [34]. Thus It is estimated that the hospital facilities should have been filled with the SARS-COV-2 patients by the end of June 2020. But we observe contrary scenario as there remain vacancies in hospital beds [17]. We project that Bangladesh might see a death cases of 0.3 to 0.9 million people (Figure 4) of whom 37.08% (0.15 to 0.33 million) will be at the age above 60 years. By the end of October 2020, the number of deaths is estimated to be 10,000 if there is no implementation of effective pandemic control methods.

**Fig. 4.**
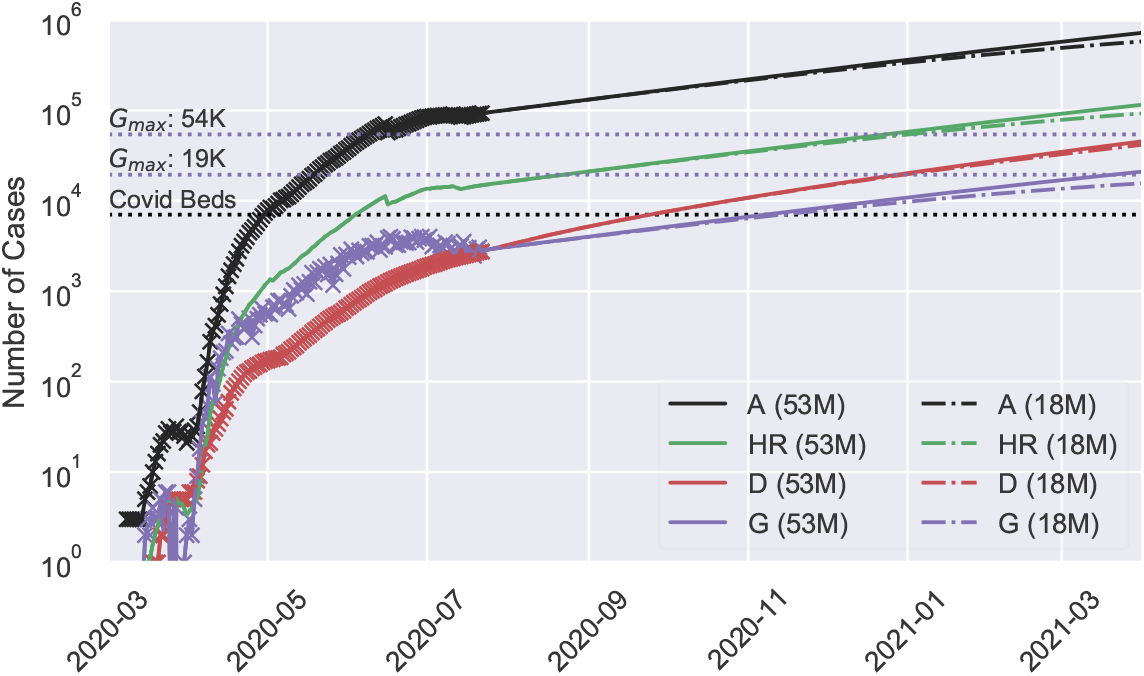
The dynamics of the active infected cases (A), corresponding hospital requirement (HR) according to the DGHS guidelines [32], deaths (D), susceptible population (S), and daily growth cases (G) due to the SARS-COV-2 in Bangladesh has been estimated for the initial susceptible population of 17.7 million (dash-dotted) and 53.0 million (solid line), respectively. The horizontal black dotted line represents the available hospital facility (7,034) for the SARSCOV-2 patient in Bangladesh. The confirmed, recovered and death cases for the COVID-19 in Bangladesh have been taken from *John-Hopkins University Database* [33] for the analysis.

### 3.3 Estimation of Case Fatality Rate (CFR)

In epidemiology, the Case Fatality Rate (CFR) is used to measure the severity of disease during an outbreak. In general, severity of disease had been defined as the percentage of affected people died in an outbreak. However, this computation has several limitations for realizing the current situation in COVID-19 pandemic. The current definition tends to underestimate the severity as it consider the rate of deceased people among the total affected people, which consider recovered, death or active cases alike. Also, this estimation does not consider the time lag between the identification of a positive case and it’s outcome (recovery and death). This issue has been discussed in detail in the literature [35, 36, 37]. So, we argue that the computation should consider the ratio between deceased, and cases with outcome instead, defined as CFR_*adj*_ [38]. We predict that at the end of pandemic, the CFR_*adj*_ in Bangladesh will be 2.10 (black line in Figure 5).

**Fig. 5.**
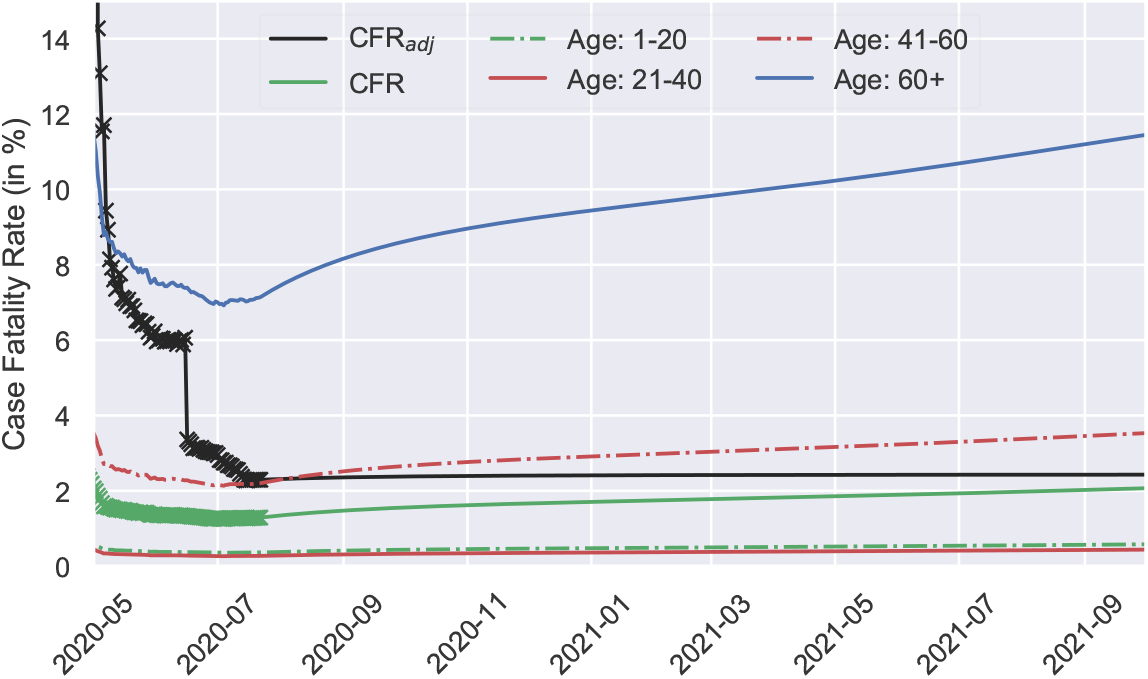
The dynamics of the CFR_*adj*_ is estimated to be about 2.1 (black solid line) at the end of the pandemic by considering the cases that have an outcome (recovered/deceased). The CFR becomes much lower (about half) if it is computed against the active cases (green dash-dotted). The cross represents the CFR corresponds to the data available. We also present the CFR for different age groups considering only the confirmed cases. The CFR at the age below 40 years, is found very low (*<* 0.6) whereas it will pass over 11.50 for the age above 60 years. In the case of age group 41–60 years, the CFR is estimated to be 3.50.

However, we are also interested in the conventional method of calculating the CFR where the ratio is considered between the deaths and confirmed cases [39, 40] as we lack sufficient data to compute the CFR_*adj*_ for different age group. We show that the CFR has been underestimated (green line in Figure 5) at the early stage of the pandemic in the traditional method which is well known in the literature [35, 36, 37]. But as both the CFR and CFR_*adj*_ converges at the end of the pandemic(black and green line in Figure 5), we choose to use the conventional method to estimate the CFR for different age groups using the available data of Bangladesh (Table 1).

The predicted CFR has a significant variation between the age below 40 and above 60+ (Figure 5). This difference illustrates the people of age 60+ as the most vulnerable group in this pandemic. We further estimate the CFR for the age group of 60+, which is above 11.5. For the age between 41 – 60, the CFR is estimated to be 3.5 while for the other age groups, the CFR is found to be less than 0.6.

## 4 Discussion

Based on our estimated population for the herd immunity, we compute the basic reproduction number to be *R*_0_ = 2.5 *±* 0.24. Therefore, to break the COVID-19 transmission chain and stop the disease from spreading, Bangladesh requires around 60% of its population to be infected to attain the herd immunity according to the existing method [41]. But our adjustment shows that if approximately 90% of the working age group 21–40 get affected by COVID-19 in case of herd immunity then we can expect a total 31% people will be affected in the whole country (presented in Table 2).

As we study the policy to contain the COVID-19 crisis in other countries, we find that Taiwan was able to manage the outbreak without imposing any lockdown [42], whereas Sweden refused to implement the lockdown claiming it as unnecessary for the nation. Nevertheless, several studies demonstrate the major role of lockdown to attenuate the contagion, though the negative impact of lockdown on national economies was not mentioned in those reports [43, 44]. Therefore, it is still a debate regarding health versus the economy for many countries, especially for a lower-middle-income country like Bangladesh. In the current state of affairs, herd immunity seems to be inevitable for COVID-19 pandemic in Bangladesh. However it might come at the expense of deaths among the vulnerable population. In this work, we analyzed this crucial trade-off in a systematic approach and synthesized a set of observations.

1. *Rapid Antibody/Antigen Testing Policy:* In order to attenuate the community transmission in Bangladesh, large scale testing strategy should be taken into account by Government policy makers. Since, Bangladesh has limited capacity for RT-PCR based COVID-19 testing, we propose low-cost rapid antibody/antigen test as soon as possible. It takes about 20–30 minutes for antibody test and around 30 minutes for antigen test [45]. The test results can also be made available to the corresponding health professional instantly. Whereas, RT-PCR based test requires 6–8 hours to be completed in the lab. In reality, it takes a few days to be made available to the corresponding health professional due to the limited lab capacity and increased number of patients. So, we suggest that each person with COVID-19 symptoms should go through an antibody/antigen test, and in case of negative results, the person should participate in an RTPCR based test for confirmation. In addition, if we can administer the rapid antibody/antigen test in the highly affected areas for everyone to get an estimation of what percentage of the population already got affected by COVID-19. This estimation will greatly help to decide on systematic implementation of pandemic control methods.
2. *Improving Health Services and Awareness:* The total number of hospital beds dedicated for the treatment of COVID-19 patients is about *∼* 7, 000 [17]. According to the current estimation in the Figure 4, hospitals in Bangladesh should have been filled up by the end of June 2020. But in reality most of the hospitals remain vacant [17]. One of the major reasons of this situation is the lack of proper health treatment for COVID-19 patients. There were numerous news of mistreatment of hospital patients followed by deaths, because of which people are now treating themselves at home. Another issue is the social stigma currently inflicted by the COVID-19 crisis. As COVID-19 patients are considered to be cursed, their entire families are being socially rejected throughout the country. As a result, COVID-19 patients like to keep themselves silent rather than seeking treatment. Hence, we need to communicate the cause of COVID-19 and health literacy to improve the situation. In this way, the health system of Bangladesh can regain its trust to contain the crisis together with the people.
3. *Collecting Pertinent COVID-19 Cases Data:* As Bangladesh is going through health crisis due to COVID-19 for the first time, we need to understand what policies can be useful for a recovery from this deadly pandemic. Countries that already became successful to contain the pandemic showed that data driven approach can be very helpful in this regard. Thus, we want to focus on collecting the COVID-19 patient data for every single patient from all over the country. We suggest that each COVID-19 test and patient data should be collected, reported on time and analyzed centrally. This will help us to predict the onset of pandemic in any area within the country as well as to design the appropriate health intervention for different location based on the need. Proper data collection will also help us to make a reliable forecast about the progression of the pandemic in the country and thus, in designing proper control methods.
4. *Policy regarding the COVID-19 vaccine allocation in Bangladesh:* The COVID-19 pandemic has given a harsh note about the dire consequences of serious diseases without vaccines. It is quite obvious that vaccines played a significant role to reduce many deaths and disability caused by preventable disease in many countries. Therefore, bio-pharmaceutical industries are trying around the clock to develop an effective, safe vaccines and the whole World is pinning its hope on COVID-19 vaccine to stop the current outbreak. Unfortunately, this is not the end of puzzle as there will be new challenges once the vaccine is available in the country. For instance, there needs to be a fair and equitable process to determine the vaccine allocation in a densely populated country like Bangladesh. Moreover, the policy makers need to decide who would get the privilege to be vaccinated first in a community. One may think whether the elders (age group 60+) should be higher up in the priority list or whether the kids should get the vaccine before the rest of the population. Since we show that age group 21–40 is the most exposed group for COVID-19 due to their social responsibilities in Bangladesh, the policy makers might consider them on the top of the list to save the socioeconomic structure of the country.

While designing the study and performing the experiment, we faced several challenges, such as, the limitation of enough COVID-19 tests restricted our understanding of actual scenario in the country, not having the information of COVID-19 test reports in a timely manner from every RT-PCR lab facilities for which making a reliable forecast became very difficult, and unavailability of the dataset in a standard format from the DGHS and IEDCR made our data analysis task very difficult. To explain the difficulties, we include a few examples in this manuscript.

For instance, if a person from a family becomes COVID-19 positive, there is a high probability that the whole family might turn out to be positive. But, due to the limited number of test capacity, the infected number reported only for a single member from that family. Moreover, it has not been possible to track each deceased person who died with COVID-19 symptoms. Again, as the COVID-19 death reports does not mention the comorbidities, it is impossible to understand the actual cause of each fatality. In addition to these, the government considered a person as recovered if that person resulted in two consecutive negative COVID-19 tests. But due to negligence, a considerable amount of the COVID-19 patients did not take the second tests. This resulted inaccurate number of recovered cases, which the government later adjusted (Figure 4 and 5). Therefore, it has been very difficult to adjust the SIRD model parameters to represent the pandemic in Bangladesh accurately. However, we are optimistic that our data-driven mathematical modeling can be used for the policy recommendations and controlling the disease transmission for the current and future outbreaks in Bangladesh.

## 5 Conclusions

Evaluation of any pandemic trend is of great importance for a nation and for its policy makers as it can guide the community towards the use of basic resources in an efficient way to mitigate the outbreak. In this manuscript, we examine the COVID-19 progression in Bangladesh by using mathematical modeling and investigate whether the current pandemic trend is destined for the herd immunity. We estimate that 17.7 to 53 million which is about 10% to 31% of the total population need to be infected by COVID-19 to achieve herd immunity in Bangladesh. In a nutshell, our mathematical approach project that herd immunity can be accomplished with less number of infected people than previously estimated [46].

We further analyzed the efficacy of past lockdown in Bangladesh and found out a similar trend in COVID-19 growth rate despite lifting the countrywide lockdown in the begining of June, 2020. The instantaneous observation of the pandemic in Bangladesh, with time-varying reproduction number (*R_t_*), is found to be independent of the implemented various control methods. Thus, continued decrease of *R_t_* does not articulate the actual scenario of the pandemic in Bangladesh. The contraction is clearly visible in the Figure 1, where we see that test positivity rate is increasing over time as opposed to the decrease of *R_t_* as presented in Figure 2. Moreover, we also present that the mobility of people in Bangladesh is increasing over time as opposed to the decreasing *R_t_* and increasing doubling time (Figure 2). So it is clearly visible that the testing capacity in Bangladesh needs to be ramped up. Also, COVID-19 test reporting system needs to be updated in a daily basis with higher precision.

We conclude that, it is debatable as though the country-wide lockdown and current zoning methods have been able to control the surge of Coronavirus infection in Bangladesh. It has been very difficult to interpret the current situation based on the COVID-19 test data as we find that total test count has fallen down despite the fact that test positivity rate is increasing. According to our SIRD estimation, we notice that the current number of confirmed case should be increasing as both the mobility and test positivity rate are increasing. Thus, Bangladesh is digressing from the path of containing the pandemic and if such a situation continues then herd immunity would be inevitable. In conclusion, using the data driven approach and SIRD modeling, we found that approximately 53.0 million people (31% of the total population) need to be infected in order to achieve herd immunity in Bangladesh.

## Data Availability

All the data and the details of computation are available in the following link.

https://github.com/mjonyh/bd_herd_immunity

## Author Contributions

MH, MI, MA, SD made substantial contributions to the conception or design of the work. MH and MI completed the relevant studies and created plots related to the SIRD estimation using Kalman filter, while MA wrote the methods for *R_t_* estimation, doubling time and growth rate computation. SD contributes to estimate the susceptible population, while DM critically revised the manuscript. All the authors met regularly to discuss the outcome of each experiment. The data were acquired from the IEDCR website and each of the results were cross validated by at least two of the three authors. All authors are responsible for acquiring, analysing, and interpreting the data for this article. MH and MI prepared the final draft. MA critically revised the Article for important intellectual content. All the authors approved the version to be published.

## Acknowledgments

We acknowledge the contribution of http://www.Pipilika.com software development team for collecting the daily data of total COVID-19 tests, total positive cases, and total deaths in Bangladesh.

## Code availability

The code relevant to this research is hosted on the Github on the following link: https://github.com/mjonyh/bd_herd_immunity.git

## Supplementary Material

This manuscript is accompanied with a supplementary material containing method details.

